# Uncovering the shared genetic contributors to primary and secondary hypertension using whole genome sequencing in a national disease cohort

**DOI:** 10.1101/2024.11.20.24317623

**Authors:** Megha Manoj, Omid Sadeghi-Alavijeh, Daniel P. Gale

**Affiliations:** Centre for Kidney and Bladder Health, University College London, London, NW3 2PF

## Abstract

**Introduction:** Extreme early-onset hypertension (EEHTN) defines a cohort of patients with a persistent blood pressure above 160/100mmHg under 30 years of age. This cohort is at heightened risk of complications and often undergo a diagnostic odyssey. We used Whole Genome Sequencing (WGS) data provided by the 100,000 Genomes Project (100KGP), to quantify the genetic contributors to EEHTN and to ascertain the diagnostic utility of WGS.

**Methods:** We performed sequencing-based genome-wide association studies (GWAS) in 901 unrelated EEHTN cases and 20852 ancestry matched unaffected controls. The analysis was inclusive of individuals with diverse genetic ancestry. Enrichment of common, low-frequency minor allele frequency (MAF) > 0.1% and rare (MAF < 0.1%) single-nucleotide variant (SNV), insertion/deletion variants (indels) and rare structural variant (SV) alleles on a genome-wide and per-gene basis was sought using a generalised linear mixed model approach to account for population structure. A validated polygenic risk score (PRS) for hypertension (HTN) was applied to the EEHTN cohort and a primary HTN cohort.

**Results:** Analysis of rare SNVs and indels revealed *PKD1* (*P=2.70 x 10^-13^*) as significantly associated with EEHTN. This signal was lost when we excluded those with known renal disease. 81.5% of the individuals harbouring qualifying *PKD1* variants had known cystic kidney disease (CyKD), this is replicated in the UK Biobank (UKBB). There were no common SNVs or rare SV associations with EEHTN. PRS discriminated between cases and controls (*P = 2.2x10^-16^*) but not between patients with PHTN or secondary HTN.

**Conclusions:** These findings represent a thorough examination of the genetic architecture of a national EEHTN cohort using well-controlled statistical methodology. The low diagnostic yield of WGS in this group brings into question its utility as a population level clinical tool but provides insights into EEHTN biology. The PRS findings suggest shared genetic contributors to early-onset extreme hypertension and primary hypertension ascertained at any age.

## Introduction

The global impact of hypertension (HTN) is significant, affecting approximately a quarter of the world’s population and representing the primary modifiable risk factor for cardiovascular disease and mortality (1). Traditionally, treatment efforts have prioritized individuals with the highest 10-year risk of cardiovascular events, often considering age as a significant determinant (2). However, HTN at a young age substantially elevates the risk of cardiovascular events later in life (3). The presence of HTN during early adulthood contributes to the premature onset of coronary heart disease, heart failure, stroke and transient ischaemic attacks. Notably, blood pressure tends to track strongly with age, meaning that elevated blood pressure in youth is likely to persist into later life (4). Studies have linked baseline blood pressure in young adults to cardiovascular mortality in follow-up assessments (5).

The evidence highlights the significance of early-life risk factors in determining long-term health outcomes and suggests that delaying the consideration of cardiovascular health until middle age may not be appropriate. With this there has been great interest in early diagnosis of HTN particularly those secondary cases where a fixed and potentially treatable cause can be found (6).

Investigating young adults for secondary causes of HTN is strongly advocated by most international guidelines due to the higher likelihood of diagnosing a secondary cause (7–9). At present the process of diagnosis for these patients is both expensive and invasive necessitating a large battery of tests at great cost (10).

In recent years there has been great interest in understanding the genetic contributors to primary hypertension (PHTN) (11). The high prevalence of HTN and its associated morbidity present a clear justification for genetic studies. Multiple genome-wide association studies (GWAS) had implicated over 50 single nucleotide variants (SNVs) associated with PHTN, with the estimated heritability from these being roughly 2%, despite family studies estimating between 30-50%, with most of the loci yet to have their functional consequences fully understood (12)

Monogenic syndromes and low frequency variants whilst rarer, have been better characterised functionally and have provided significant biological insights with >25 genes implicated, helping to elucidate the pathways behind salt retention and underlying the role of the kidneys and adrenal glands in blood pressure regulation. Most of the rare variant work has been conducted using linkage analysis with a few exome sequencing projects yielding mixed results probably due to low sample sizes (13) (14). Whole genome sequencing (WGS) provides more uniform coverage and avoids capture bias, resulting in increased sensitivity to detect structural variants (SVs) and rare SNVs compared with exome (or capture-based) sequencing platforms, even withing regions targeted by the latter (15, 16). In addition, WGS benefits from economised of scale and the associated costs have dropped dramatically over the past decade (17). However, no previous case-control studies have looked at the utility of WGS in HTN on a large scale.

Anticipated enrichment for secondary and monogenic causes of HTN in younger cohorts with HTN was the justification to include an “extreme early-onset HTN” (EEHTN) cohort in the 100,000 genomes project (100KGP). EEHTN is defined as a blood pressure in an adult > 160/100mmHg in clinic and an average blood pressure of 150/95 on ambulatory blood pressure monitoring occurring below the age of 30 (excluding patients with primary hyperaldosteronism, phaeochromocytoma, Cushing’s syndrome and hyper/hypothyroidism).

In this study we utilised WGS to perform association studies across the full spectrum of genetic variation in 901 EEHTN cases and 20411 ancestry matched controls. We find an enrichment for rare variants in *PKD1*, which is lost when removing patients with a known renal diagnosis, and a similarity between EEHTN and PHTN at a common variant level.

## Methods

### The 100,000 Genomes Project

The 100KGP is one of the largest disease-based sequencing initiatives in the world in which WGS data from large numbers of National Health Service (NHS) patients with rare diseases and cancer, and their relatives, have been generated (18). Key strengths of this dataset with respect to the study of rare diseases are that all germline samples are processed and analysed using a shared pipeline and that sequencing data is available for many individuals without the phenotype under study, drawn from the same population. This allows for robust control of technical artefacts, allele frequency and variant burden in the population, in contrast to previous sequencing studies.

Recruitment to the 100KGP is via a network of 13 NHS Genomic Medicine Centres (GMCs) and includes collections of phenotype data hierarchically encoded using Human Phenotype Ontology (HPO) codes (19), facilitating computerised analysis of clinical features. EEHTN patients were recruited to the project if they met the following criteria:

1. A blood pressure in an adult > 160/100mmHg in clinic and an average blood pressure of 150/95mmHg on ambulatory blood pressure monitoring occurring below the age of 30

Participants were excluded if they were found to have primary hyperaldosteronism, phaeochromocytoma, Cushing’s syndrome and hyper/hypothyroidism.

All the recruited EEHTN cases underwent assessment via the clinical interpretation arm of the 100KGP to obtain a molecular diagnosis via PanelApp (20). This involves looking for known causative variants or those that are predicted to be damaging in the set of 27 genes, listed on PanelApp, that have been found to be associated with secondary causes of HTN. Following on from this, a multidisciplinary panel conducted a review and the American College of Molecular Genetics and Genomics (ACMG) criteria (21) were applied for determining pathogenicity. This information was then used to categorise cases into ‘solved’, ‘partially solved’ or ‘unsolved’. All this data was then stored on the Genomics England servers to be accessed through LabKey (22), a data integration platform, using R (23).

Ethical approval for the 100KGP was granted by the Research Ethics Committee for East of England - Cambridge South (REC Ref 14/EE/1112).

### Cohort creation

All the cohorts used were unrelated and ancestry matched in order to maximise power and improve inclusivity without affecting genomic inflation (this is detailed in full in supplementary methods).

We defined three cohorts for downstream association analysis:

1. An EEHTN cohort made up of those probands recruited to 100KGP as defined above. After the full process of removing related patients, ancestry matching and filtering there were 179 cases.
2. A hospital episode statistic (HES) generated cohort generated by finding individuals for whom the earliest recorded hypertensive event (as defined by HES coding) occurred at or under the age of 30. This was combined with the first cohort and labelled the HES-EEHTN group. After the full process of removing related patients, ancestry matching and filtering there were 901 cases.
3. The same HES-EEHTN cohort but with all causes of renal disease removed due to the 100KGP being enriched for renal disease probands, labelled the RR-EEHTN. After the full process of removing related patients, ancestry matching and filtering there were 449 cases.

All cohorts were generated by querying stored recruitments and coded phenotype data via the Labkey interface in R (22, 23).

### DNA preparation and clinical pipeline

DNA was extracted and WGS was performed by Genomics England (24), with the output being fed into the 100KGP clinical pipeline as previously described (25).

### Association analyses

#### Rare variant analysis

We used a collapsing approach Scalable and Accurate Implementation of Generalized mixed model (SAIGE)-GENE when conducting the rare variant analysis, as this approach increases power and reduces type I error which occurs due to the substantial number of rare variants each individual harbours (26). Rare variant collapsing analysis works on the principle that those variants selected for, that form a functional unit for testing, are likely to be causative (27). Thus, in all three cohorts, we chose to include SNVs and insertion/deletion variants (indels) which were rare, with a MAF < 0.001 (0.1%) as seen in the gnomAD database (28) or not seen in gnomAD at all, with a Combined Annotation-Dependent Depletion (CADD) score, that measures deleteriousness (29) over 20 (corresponding to the top 1% of all predicted deleterious variants in the genome) and that included missense, in-frame insertion, in-frame deletion, start loss, stop gain, frameshift, splice donor or splice acceptor variations. Of note we also included missing CADD scores as indels do not have a CADD score annotation. We iterated these variant annotations into different filters that were run concurrently, these were:

⇒ Loss of Function (LoF): filter taking only variants predicted to cause protein truncation as per citation (30).
⇒ Missense+: missense variants and variants that are listed on Ensembl to be of a higher impact than missense, including frameshift variants and splice donor variants (31).

The results of a rare variant analysis are presented on a gene-based Manhattan style plot showing the -log_10_ of the P values of individual rare variants tested for (y-axis), and the position of the gene on the chromosome (x-axis). Each point represents a gene made up of the qualifying variants as per the relevant filter, with MAF < 0.001 (0.1%) in gnomAD. As we used different annotations when testing for rare variants i.e. LoF and missense+, separate graphs have been generated showing each of these filters. These were plotted in R using the qqman package (32).

Phenotype information for patients with qualifying variants in the most significant genes was extracted from labkey using R.

### Common variant analysis

SAIGE, which utilises a generalised linear mixed model, was used for common variant analysis (33). In this analysis the top ten principal components and sex were used as covariates, and SNVs and indels with MAF > 0.01 (1%) were included.

Manhattan and Quantile-Quantile (Q-Q) plots were generated using the R package qqman.

### SV analysis

Exon crossing SV and Copy Number Variants (CNVs) in genes known to cause secondary HTN as set out in PanelApp (20) were counted in cases versus controls.

This was done using the SV analysis pipeline developed by the Genomics England team (see Data Availability). CANVAS (v1.3.1) (34) and MANTA (v0.28.0) (35) were used to determine copy number (>10 kilobases) and find SVs of size greater than 50 base pairs respectively. Variants were separated by the specific type of SVs (INV: inversion. DEL: deletion. BND: translocation break-end. DUP: duplication. LOH: loss of heterozygosity. INS: insertion). For inversions, the gnomAD SV calling protocol was followed whereby only inversions where the breakpoints fell within the gene or those that were exon spanning were counted (36). The human genome contains many whole gene spanning inversions, and their biological relevance is unascertained (35). The number of variants per gene per subtype of SV were compared in cases and controls using a two tailed Fisher’s exact test (37) in R.

### Polygenic Risk Scoring

A polygenic risk score (PRS) was applied from a pre-validated PRS of HTN (38), which predominantly included patients with PHTN. The PRS was applied to the EEHTN cohort (179 cases, 20411 controls), the HES-EEHTN cohort (901 cases, 20852 controls), the RR-EEHTN cohort (449 cases, 20852 controls) and the PHTN cohort (7923 cases, ∼20000 controls). The PRS included 186,135 markers derived from the UK Biobank (UKBB) using LDpred (39). This was lifted over using the USC Liftover tool (40) from build 37 to 38 and imported into the 100KGP where the scoring was performed using the “score” command in PLINK2 (41). To test the significance between the PRS of cases and controls we applied a Kruskal-Wallis test in R. All plotting was performed with ggplot2 in R (42).

### P value adjustment

For rare variant and SV/CNV collapsing analyses, the Bonferroni adjusted exome-wide significance threshold was defined on a per gene level as PLJ= (0.05/19,000)LJ≈ *2.6*!J*×*!J*10^−6^* (43). For common variant analysis the Bonferroni adjusted significance threshold was P = *5 x 10^-8^*.

### Power Calculation

Power was calculated using the genpwr tool in R (44). Under the assumption of a case rate of 4% of the population having a secondary cause of HTN (45) a range of powers were calculated across MAFs. Assuming a MAF <0.001 we found we needed a cohort of between 15343 and 30405741 to achieve a power of at least 0.8 (80%) for an assumed P value of <0.05 (after multiple testing correction) for a dominant and recessive model respectively.

## Results

### The EEHTN cohort

There are 200 probands recruited with EEHTN in 100KGP, from which 179 cases were derived for the EEHTN cohort accounting for the genome build alignment and ancestry matching. Table 1 highlights the demographic information of the recruited cohort.

**Table 1.**
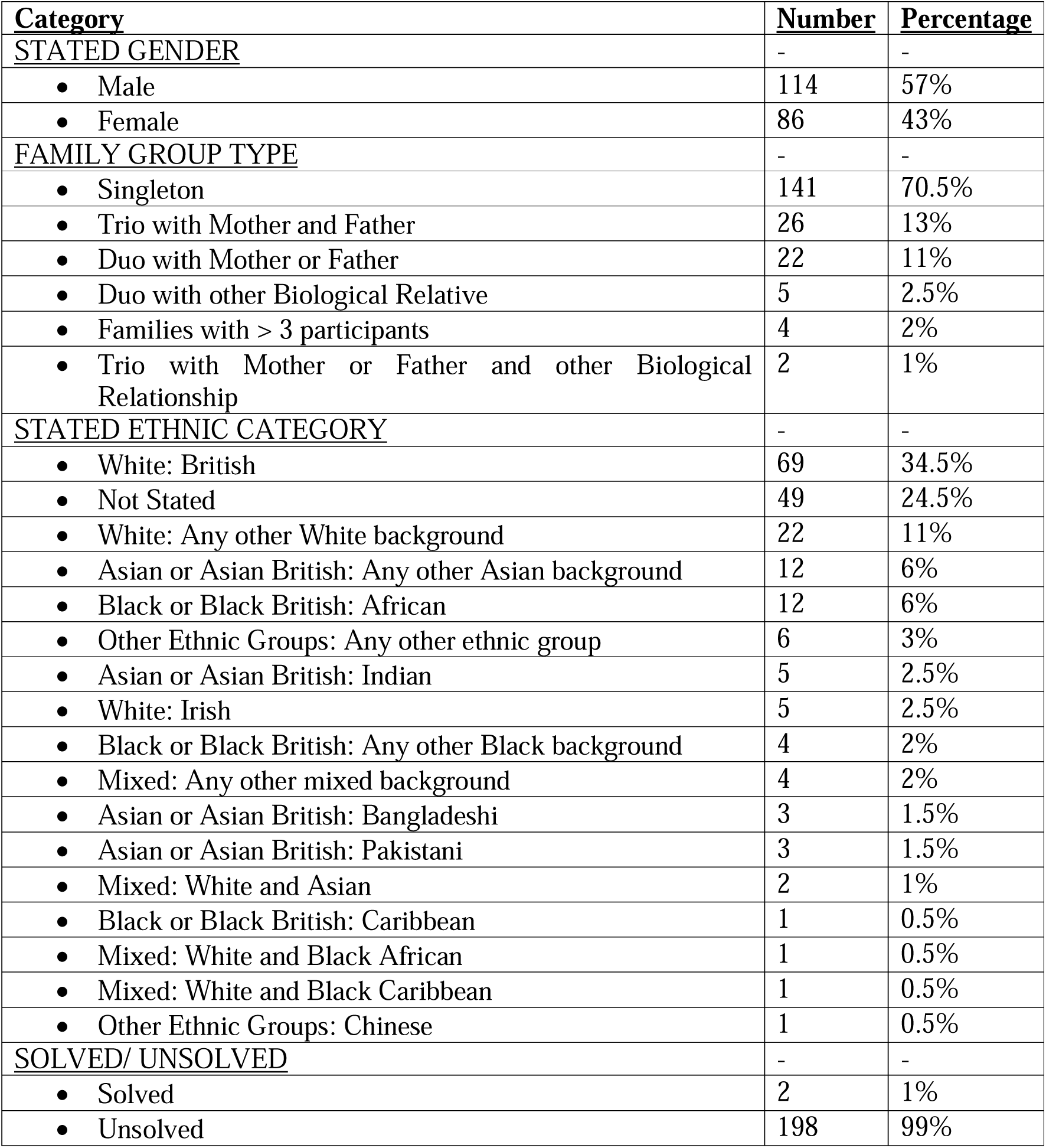
Table showing EEHTN demographics.

| **<u>Category</u>** | **<u>Number</u>** | **<u>Percentage</u>** |
| --- | --- | --- |
| **<u>STATED GENDER</u>** | - | - |
| • Male | 114 | 57% |
| • Female | 86 | 43% |
| **<u>FAMILY GROUP TYPE</u>** | - | - |
| • Singleton | 141 | 70.5% |
| • Trio with Mother and Father | 26 | 13% |
| • Duo with Mother or Father | 22 | 11% |
| • Duo with other Biological Relative | 5 | 2.5% |
| • Families with > 3 participants | 4 | 2% |
| • Trio with Mother or Father and other Biological Relationship | 2 | 1% |
| **<u>STATED ETHNIC CATEGORY</u>** | - | - |
| • White: British | 69 | 34.5% |
| • Not Stated | 49 | 24.5% |
| • White: Any other White background | 22 | 11% |
| • Asian or Asian British: Any other Asian background | 12 | 6% |
| • Black or Black British: African | 12 | 6% |
| • Other Ethnic Groups: Any other ethnic group | 6 | 3% |
| • Asian or Asian British: Indian | 5 | 2.5% |
| • White: Irish | 5 | 2.5% |
| • Black or Black British: Any other Black background | 4 | 2% |
| • Mixed: Any other mixed background | 4 | 2% |
| • Asian or Asian British: Bangladeshi | 3 | 1.5% |
| • Asian or Asian British: Pakistani | 3 | 1.5% |
| • Mixed: White and Asian | 2 | 1% |
| • Black or Black British: Caribbean | 1 | 0.5% |
| • Mixed: White and Black African | 1 | 0.5% |
| • Mixed: White and Black Caribbean | 1 | 0.5% |
| • Other Ethnic Groups: Chinese | 1 | 0.5% |
| **<u>SOLVED/ UNSOLVED</u>** | - | - |
| • Solved | 2 | 1% |
| • Unsolved | 198 | 99% |

### Collapsing rare variant analysis highlights Autosomal Dominant Polycystic Kidney Disease (ADPKD)-*PKD1* as an important driver of EEHTN

In the HES cohort (n=901) there was significant enrichment of LoF variants in the *PKD1* gene (*P=2.70x10^-13^*) compared to controls (n=20852) (Fig. 1). There was no enrichment in the missense+ annotation (Fig.S7)

**Fig. 1.**
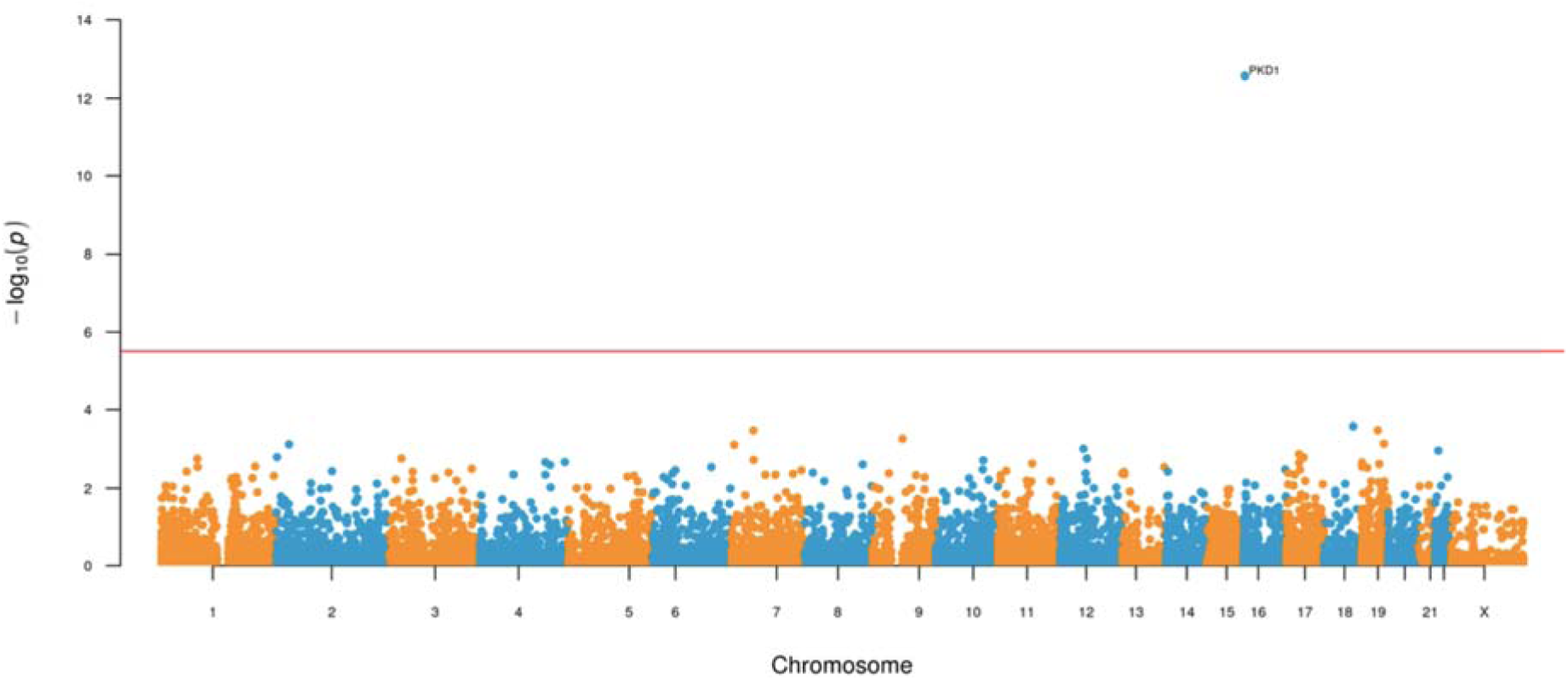
Manhattan style plot showing the results of the rare variant analysis: Manhattan style plot showing the results of the rare variant analysis, each point represents a gene made up of qualifying variants. Variants were included if they were predicted to be LoF. There is enrichment *(P=2.70 x 10^-13^)* of LoF rare variants in *PKD1* affecting the EEHTN phenotype in this cohort of 901 cases v 20852 controls.

In both the EEHTN cohort (n=179) and the RR-EEHTN cohort (n=449) there was no significant genome-wide enrichment in any single gene compared to controls (EEHTN [n=20411], RR-EEHTN [n=20852]) (Figures S3,S4,S5 and S6) in either the LoF or missense+ annotations.

Further analysis of the 27 cases whose variants contributed to the *PKD1* association revealed that all the variants were heterozygous being made up of 13 frameshift variants, 10 stop gain variants, 2 splice donor variants and 2 missense variants.

The recruited diseases for the 27 patients were Congenital Anomalies of the Kidney and Urinary Tract (CAKUT) (1 patient), Cystic Kidney Disease (CyKD) (22 patients), Epilepsy (2 patients), syndromic congenital heart disease (1 patient), unexplained kidney failure in young people (1 patient).

13 out of 27 patients were solved, meaning their clinical team had been given an explaining molecular diagnosis. 16 patients had a positive family history and 2 patients had consanguineous parents (Table 2).

**Table 2.** Table summarising information about percentages and numbers of patients that make up the *PKD1* hit that are ‘solved’ (meaning they have received a genetic/molecular diagnosis), with consanguineous parents and that have family history of the condition.

| **<u>Diagnosis</u>** | **<u>Percentage solved</u>** | **<u>Percentage positive for consanguinity</u>** | **<u>Percentage with family history</u>** |
| --- | --- | --- | --- |
| CAKUT | 100% (1/1) | 0% (0/1) | 100% (1/1) |
| CyKD | 54.5% (12/22) | 9.1% (2/22) | 68.2% (15/22) |
| Epilepsy plus other features | 0% (0/2) | 0% (0/2) | 0% (0/2) |
| Syndromic congenital heart disease | 0% (0/1) | 0% (0/1) | 0% (0/1) |
| Unexplained kidney failure in young people | 0% (0/1) | 0% (0/1) | 0% (0/1) |

The top 10 recorded HPO codes for this cohort are shown in Fig.S14. There is a clear trend towards polycystic kidney disease (PKD) codes.

### PRS

Across all cohorts we found statistically significant differences between cases and controls in the HTN PRS (EEHTN: *P=7.25x10^-10^*, HES: *P=1.00x10^-15^*, RR-EEHTN: *P=5.84x10^-6^*, PHTN: *P= p value not known*). In all cohorts the cases had a higher PRS than controls as shown in Figure 2. There was no significant difference in the PRS between the primary and EEHTN cohorts.

**Fig. 2.**
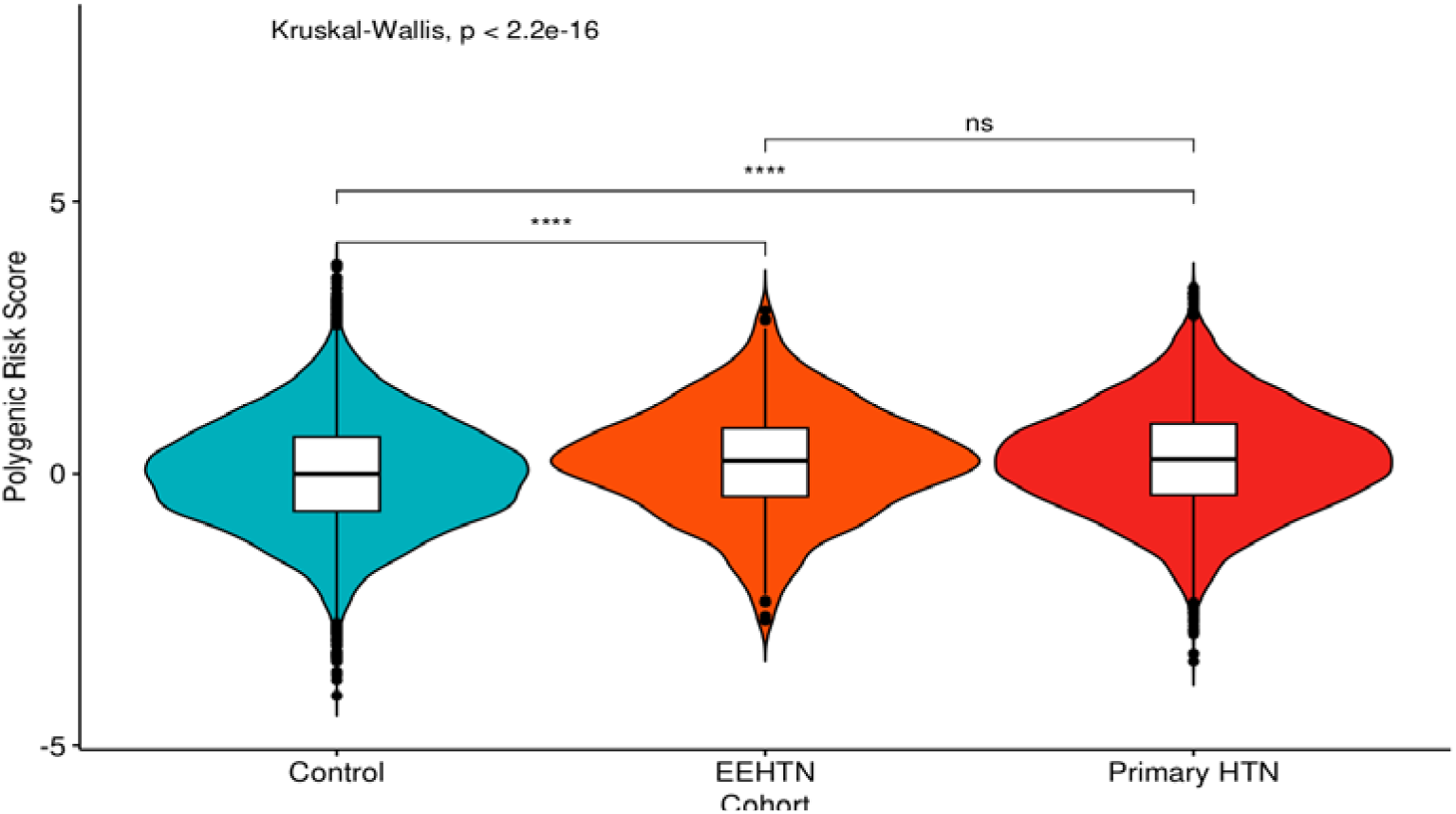
Violin and boxplot comparing Polygenic Risk Score distribution across HTN cohorts: Violin and boxplot showing the PRS distributions between controls (without PHTN or secondary HTN), EEHTN cases and cases with PHTN. The means of the three PRS were compared with a Kruskal-Wallis test (*P=2.2x10^-16^*) with the signal being driven by the difference between unsolved cases and controls. ****= statistical significance, ns= no significant difference.

### GWAS

Across all three cohorts (EEHTN [n=179], HES-EEHTN [n=901], RR-EEHTN [n=449]) there was no significant genome-wide associations (Figures S3, S5 and S7). All three cohorts were well controlled for as per their lambda (Figures S4, S6 and S8).

### SV analysis

There was no enrichment in any SV at a case control level. Of note there was a single case with a *WNK1* exon spanning CNV loss (Chr12:765990-880472), which was not seen in the ClinVar database (46), however there was a comparable CNV loss seen in another individual that was linked to Pseudohypoaldosteronism type II (ClinVar: 5161).

## Discussion

Rare and common variant analysis using the EEHTN, HES-EEHTN and RR-EEHTN cohorts found no genetic associations with secondary HTN genome-wide, bar statistically significant enrichment of rare variants in the *PKD1* gene in the HES-EEHTN cohort, reflecting the power of this cohort to detect genetic associations. This finding and our approach is validated by a UKBB rare variant analysis study for secondary HTN, analysing 281,104 individuals, where there is a similar finding of no known causes of secondary HTN bar *PKD1* (47).

### Evaluation of WGS

As mentioned previously, diagnosing patients with EEHTN is a long and arduous process involving 24-hour urine collection, a myriad of blood tests and complex imaging which potentially culminate in complex adrenal vein sampling or other invasive tests (48). This delays diagnosis and can affect outcomes. WGS has been looked at as a potential solution to these issues (49) as it is relatively quick and non-invasive, requiring a simple blood test. The clinical genomics pipeline of the 100KGP EEHTN pipeline has only yielded a 1% (2/200 participants) success rate of molecular diagnosis in the cohort thus far. If one includes the additional solved HES cohort with *PKD1* variants an additional 13 are solved. This represents an extremely low return, particularly when compared to other heterogenous cohorts such as those patients with CyKD where the diagnosis rate of WGS can approach ∼10% (50). At present it would be difficult to recommend WGS for use in an EEHTN cohort however, the key caveat would be that the recruited cohort were already clinically deplete for known causes of secondary HTN. A further study of new referrals to a secondary HTN service prior to work-up comparing the diagnosis rates between traditional clinical testing and WGS would help answer the question of WGS utility more robustly.

A key limitation of this study is the limitation of power to detect potential associations, so it is hoped that further, ideally larger, studies may be performed in the future to increase the chances of identifying new genetic contributors to EEHTN. As Genomics England is part of the UK Genomics Testing Service, it is possible that this goal could be met in the near future as more people are undergoing WGS, increasing the numbers of participants available for association analyses.

Subsetting patients according to their conditions and conducting association analyses within these smaller groups would also be a useful next step to determine whether a genetic cause can be found in people with known disease. However, a required prerequisite for this would again be an expanded cohort, as the number of participants is currently too small to have sufficient power for the association analyses after subsetting. As mentioned above, a study comparing patients undergoing WGS or the traditional diagnostic work-up for EEHTN would be the gold standard to compare methodologies.

### Limitations

Since power to detect genetic association is a function of the cohort size, and both the frequency and effect size (i.e. relative risk) of genetic variants it is unsurprising that this study did not identify previously undetected genetic contributors to HTN, which can result from multiple different pathologies. This heterogeneity renders the cohort underpowered to detect a signal for any one disease, except for *PKD1*-associated HTN, reflecting the frequency of PKD – both within this study and within the general population. The EEHTN cohort were entered into the 100KGP after they had already undergone screening for known causes of HTN making it even less likely that they would be enriched for such causes, consistent with previous literature reporting that rare variants causing HTN were too infrequent to be found on a population scale (12).

90-95% of hypertensive patients have PHTN, which is highly heterogeneous and influenced by gene-environment interactions (51). Significant SNVs were only discovered in PHTN after the cohort sizes far exceeded those available in this study (52). A limitation of GWAS is that they are likely to be underpowered to pick up all heritability caused by SNVs and indels as the significance level is around *P < 5 x 10^-8^*, after accounting for multiple testing. Increasing the sample size will help overcome this problem, as it increases the power. However, this can be difficult for a rare disease where the sufficient sample size to have adequate power might be difficult to achieve (53).

This means that, for many patients, it might be difficult to pinpoint a single gene containing rare variants, SNVs or indels, as HTN is not a single disease.

The fact that the UKBB rare variant analysis study did not find known causes of secondary HTN in its analysis of 281,104 individuals highlights the numbers that are required to find genes that enrich in secondary HTN bar *PKD1* (47). Our power calculations suggested that under a dominant model of inheritance the analyses did have sufficient power to detect a genetic signal (*PKD1*), however, this was not the case under a recessive model of inheritance. Therefore, as the model of inheritance could be variable it is likely that there was insufficient power to identify certain genes as linked to the EEHTN phenotype. This study underlines the fact that EEHTN is almost always not a monogenic disorder.

In the HES-EEHTN cohort, a statistically significant enrichment of rare variants in the *PKD1* gene (*P=2.70 x 10^-13^*) was found. The 100KGP is enriched for patients with CyKD (>1200 probands have been recruited for this disease), many of whom develop EEHTN. Encouragingly these results have been validated in an independent cohort in the UK where *PKD1* (*P=8.16 x 10^-13^*) is enriched for the secondary HTN phenotype (47).

Whilst the SV analysis yielded interesting results, novel SV calling with short read technology is fraught with difficulties and at the minimum required the use of multiple SV calling algorithms (54).

### Conclusions

In conclusion, rare variants in *PKD1* were found to have a significant association with the EEHTN phenotype, however this is an already well characterised disease and the strong signal may be due to the enrichment of renal patients in the 100KGP. No other genes with rare variants, SNVs or indels were found to have a significant relation with EEHTN. The PRS analyses suggest that most of these individuals with EEHTN have PHTN that has become clinically appreciable at an earlier age, likely owing to unascertained environmental or genetic contributors which may be detectable in future studies with the appropriate design and size to detect them. These results suggest that WGS is not clinically applicable for diagnosis of EEHTN as EEHTN is generally not a monogenic disease. However, as WGS is being conducted for more patients, further studies with expanded cohort size might provide additional insight into the genetic architecture of EEHTN.

## Non-standard Abbreviations and Acronyms

100KGP: 100,000 Genomes Project
ACMG: American College of Molecular Genetics and Genomics
ADPKD: Autosomal Dominant Polycystic Kidney Disease
AggVCF: Aggregated Variant Calling File
BND: Translocation Break-End
CADD: Combined Annotation-Dependent Depletion
CAKUT: Congenital Anomalies of the Kidney and Urinary Tract
CNV: Copy Number Variant
CyKD: Cystic Kidney Disease
DEL: Deletion
DUP: Duplication
EEHTN: Extreme early-onset hypertension
FDR: False Discovery Rate
gVCF: Genomic Variant Calling File
GMC: Genomic Medicine Centre
GWAS: Genome-Wide Association Study
HES: Hospital Episode Statistics
HPO: Human Phenotype Ontology
HTN: Hypertension
Indel: Insertion/Deletion Variant
INS: Insertion
INV: Inversion
KING: Kinship-based Inference for Genome-wide association studies
LoF: Loss of Function
LOH: Loss of Heterozygosity
MAF: Minor Allele Frequency
NHS: National Health Service
PHTN: Primary Hypertension
PKD: Polycystic Kidney Disease
PRS: Polygenic Risk Score
Q-Q plot: Quantile-Quantile plot
RR-EEHTN: Hospital Episode Statistics generated cohort without patients with known renal conditions
SAIGE: Scalable and Accurate Implementation of Generalized mixed model
SNV: Single Nucleotide Variant
SV: Structural Variant
UKBB: UK Biobank
WGS: Whole Genome Sequencing

## Data availability

General description of the 100KGP dataset:

- https://research-help.genomicsengland.co.uk/display/GERE/Research+Environment+User+Guide

Aggregated dataset of 78,195 people:

- https://research-help.genomicsengland.co.uk/pages/viewpage.action?pageId=38046780

Rare variant workflow:

- https://research-help.genomicsengland.co.uk/display/GERE/AVT+workflow+v3.x+docs

Common variant (GWAS) workflow:

- https://research-help.genomicsengland.co.uk/display/GERE/GWAS+Nextflow+Pipeline

SV analysis workflow:

- https://research-help.genomicsengland.co.uk/display/GERE/SVCNVworkflow+v1.2

## Supplemental materials

Supplementary methods

Figures S1-S14

