## Supplementary Information for "Uncovering the shared genetic contributors to primary and secondary hypertension using whole genome sequencing in a national disease cohort"

**Supplementary methods**

### **Supplementary methods 1: EEHTN cohort and controls ancestry matching, relatedness filtering and quality control**

Central to this analysis were the aggregated variant calling files (AggVCF) and PLINK (55) files generated by the central bioinformatics team at Genomics England. This consisted of WGS data from 78,195 100KGP participants (see Data Availability) aggregated into a large genomic VCF (gVCF) file chunked by chromosomal location (the aggv2 file – see Data Availability). Alongside this a corresponding set of autosomal SNVs which had been pruned for LD and with MAF > 0.01 (1%) were also provided.

On a per cohort basis we used Kinship-based Inference for Genome-wide association studies (KING) (56) to remove related cases using the PLINK aggv2 files. We then merged the cases and controls and then employed KING again, using a custom python script developed by another member of the lab, to remove relatedness between cases and controls. Using this unrelated case control cohort we generated 10 principal components in PLINK for use as covariates in the association analyses to control for population stratification (57).

In order to increase the numbers of cases and controls we used an ancestry matching strategy, as detailed in this paper (58), to find genetically similar controls who clustered around each case (Figures S1 and S2) using weighted principle components.

This resulted in a final total of 179 cases and 20411 controls in the original EEHTN cohort, 901 cases and 20852 controls in the HES-EEHTN cohort and 449 cases and 20852 controls in the RR-EEHTN cohort.

A primary HTN cohort was created by taking unrelated individuals recruited to 100KGP with a diagnosis of “essential” or “primary” HTN from their ICD-10 codes who were at least 40 years of age at the time of diagnosis, they were then depleted for any secondary cause for HTN and ancestry matched to similar controls to above leaving 7923 primary HTN cases and ~20000 controls.

**Supplementary data:**

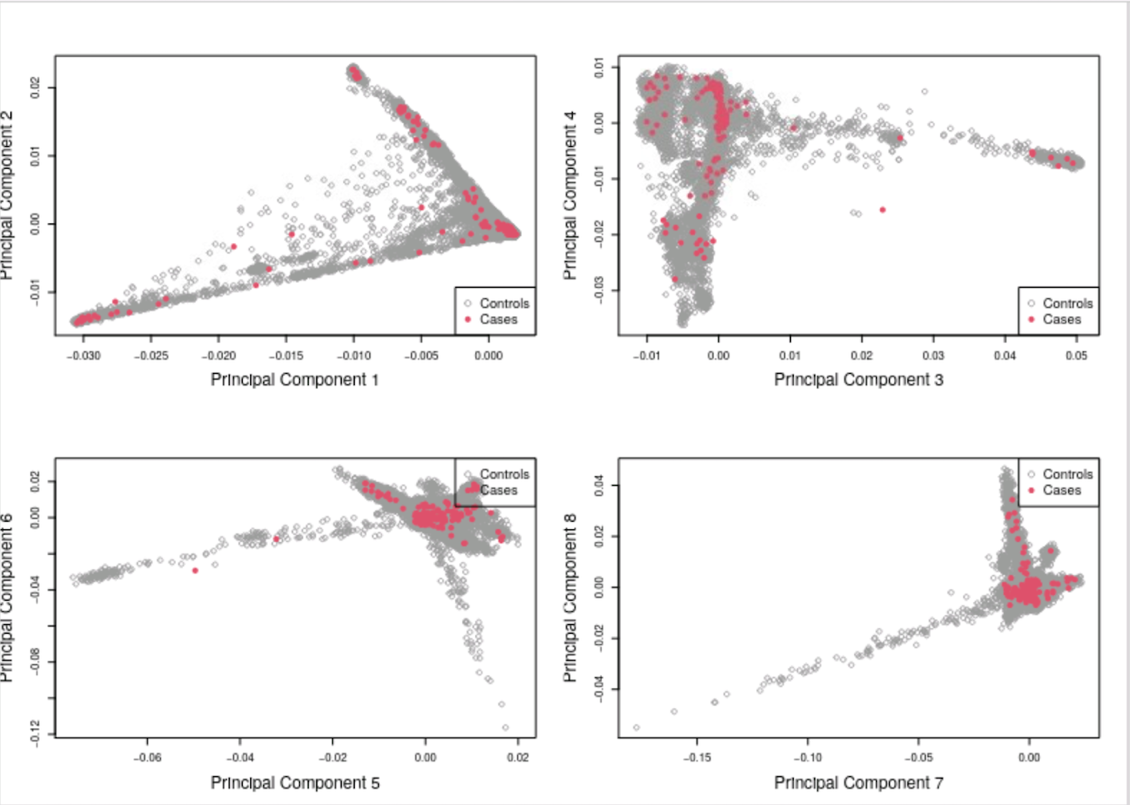

**Fig.S1a** Principal component plot for original EEHTN cohort, showing all the cases and potential controls before application of the ancestry matching algorithm.

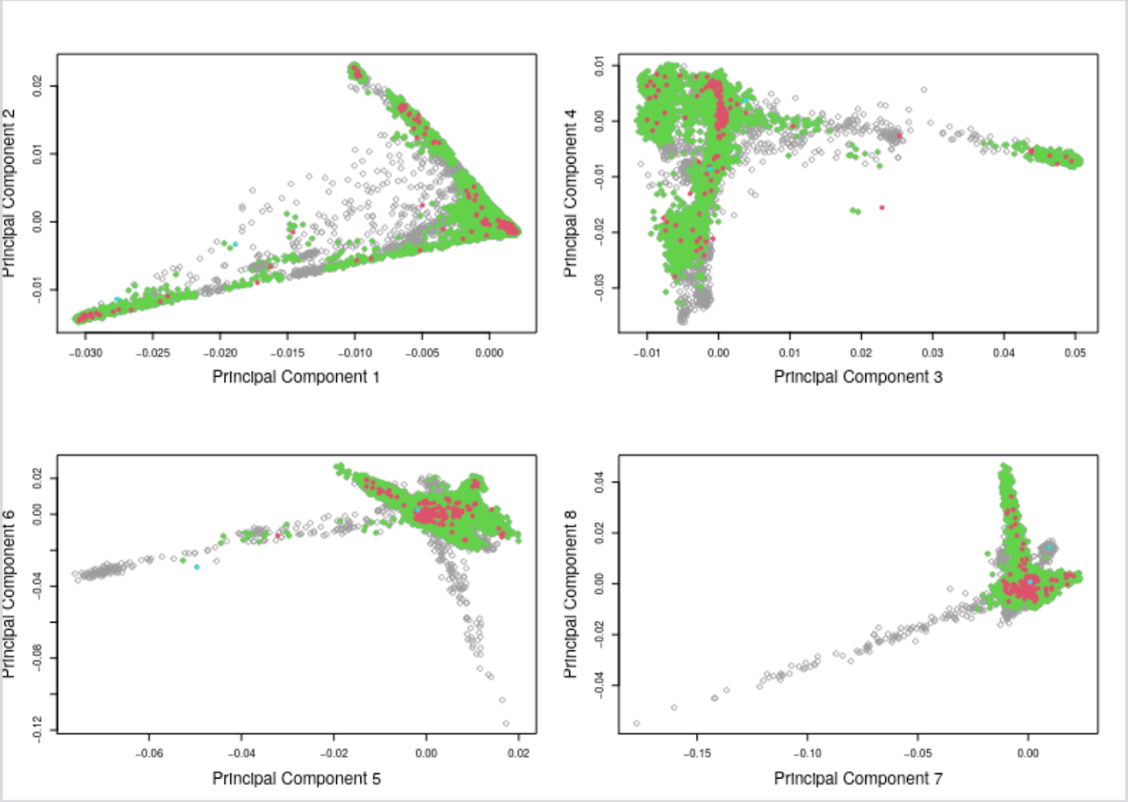

**Fig.S1b** Principal component plot for original EEHTN cohort showing all cases (red), and controls (green) matched to cases after application of ancestry matching algorithm.

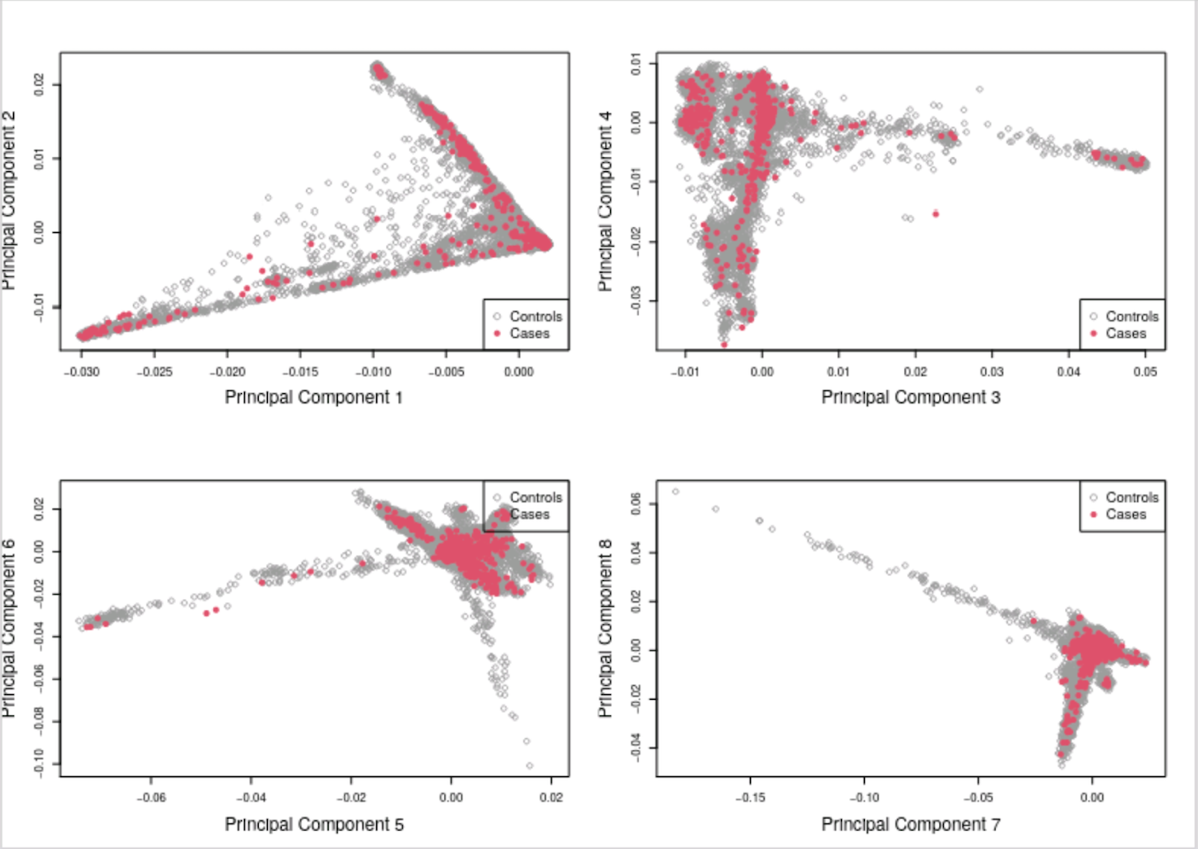

**Fig.S2a** Principal component plot for the HES generated cohort, showing the possible cases and controls before application of the ancestry matching algorithm.

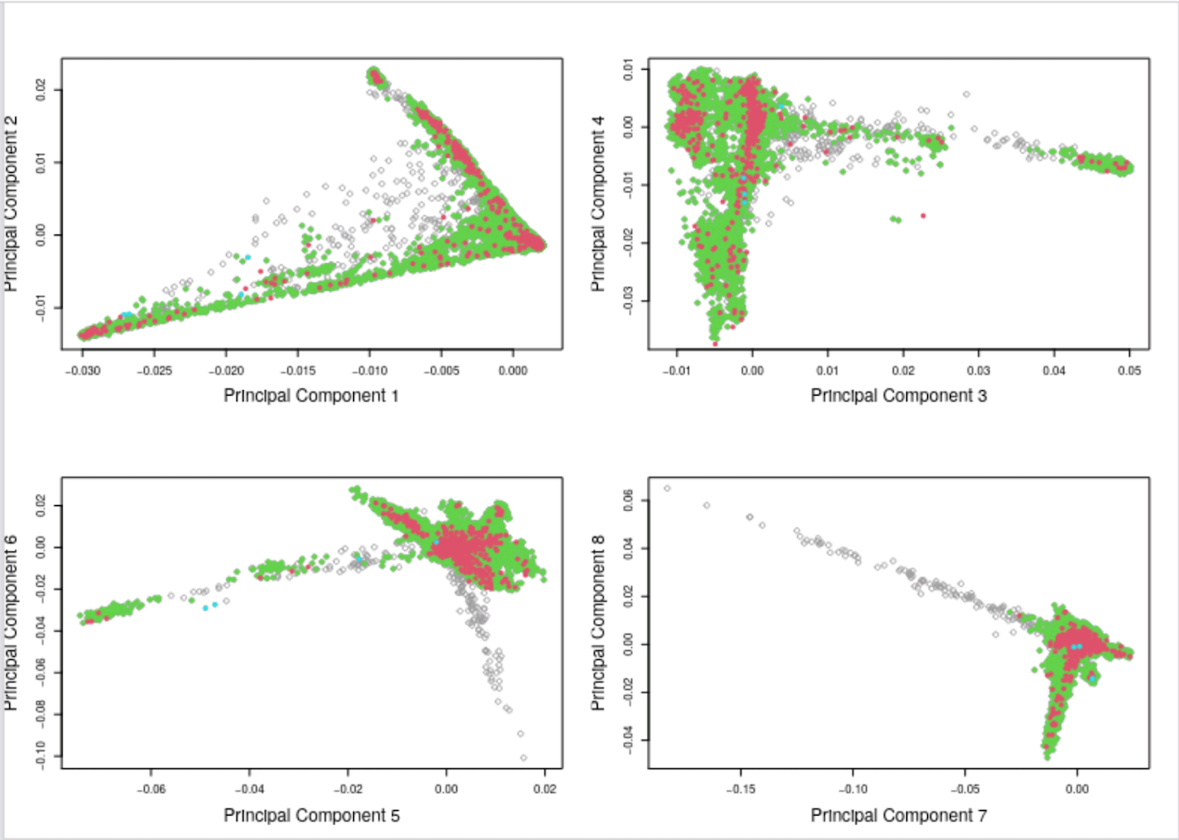

**Fig.S2b** Principal component plot for the HES-EEHTN cohort showing all cases (red), and controls (green) matched to cases after application of ancestry matching algorithm.

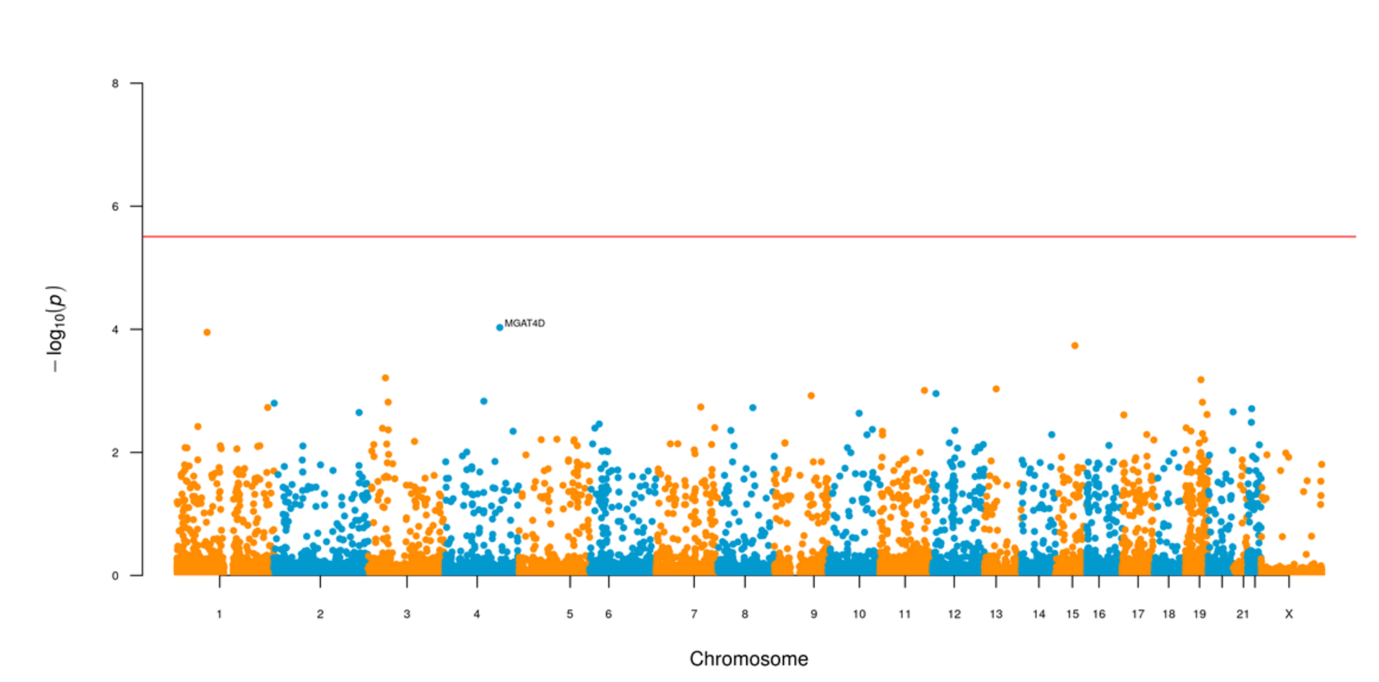

**Fig.S3** Manhattan style plot showing the results of the rare variant analysis, using gnomAD, for LoF variants with MAF < 0.001 (0.1%) in the original EEHTN cohort, with the red line representing the -log_10_ of the P value (~*2.6 x 10^-6^*) that is the threshold for genome-wide significance. There is no enrichment of any LoF rare variant affecting the EEHTN phenotype in this cohort of 179 cases v 20411 controls.

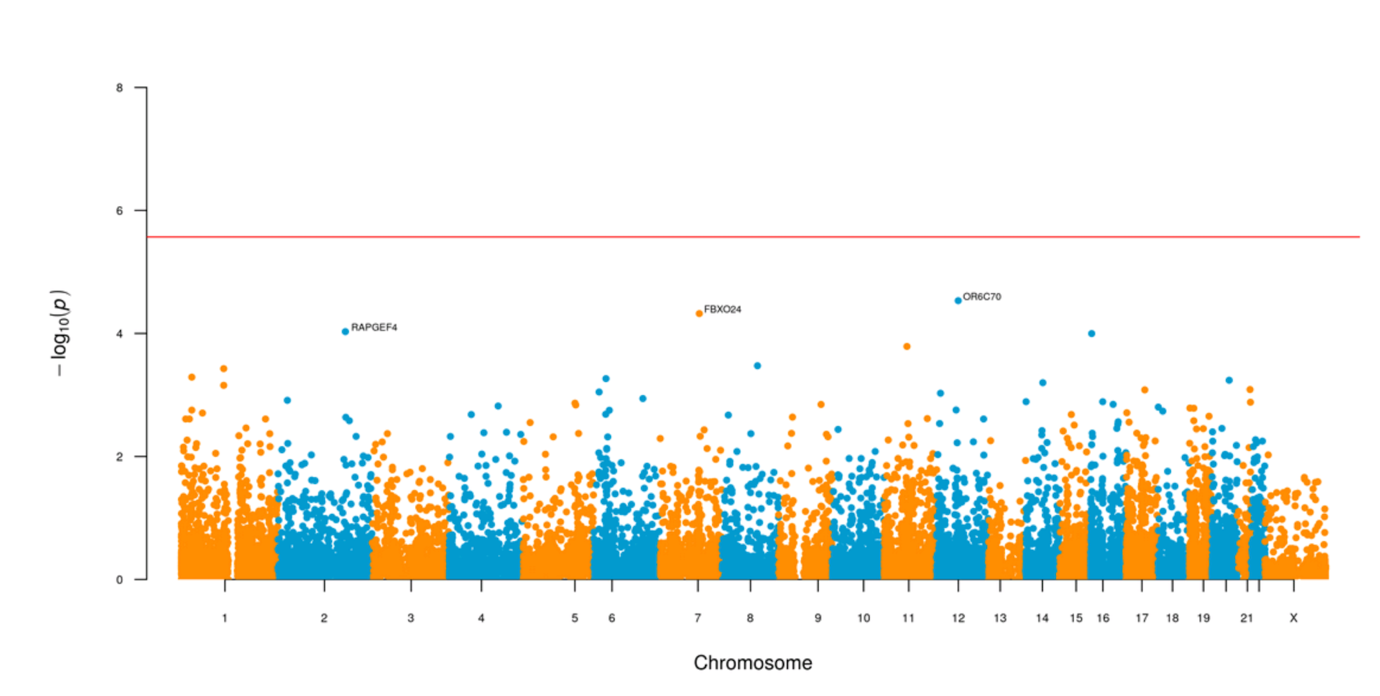

**Fig.S4** Manhattan style plot showing the results of the rare variant analysis, using gnomAD, for missense+ variants with MAF < 0.001 (0.1%) in the original EEHTN cohort, with the red line representing the -log_10_ of the P value (~*2.6 x 10^-6^*) that is the threshold for genome-wide significance. There is no enrichment of any missense+ rare variant affecting the EEHTN phenotype in this cohort of 179 cases v 20411 controls.

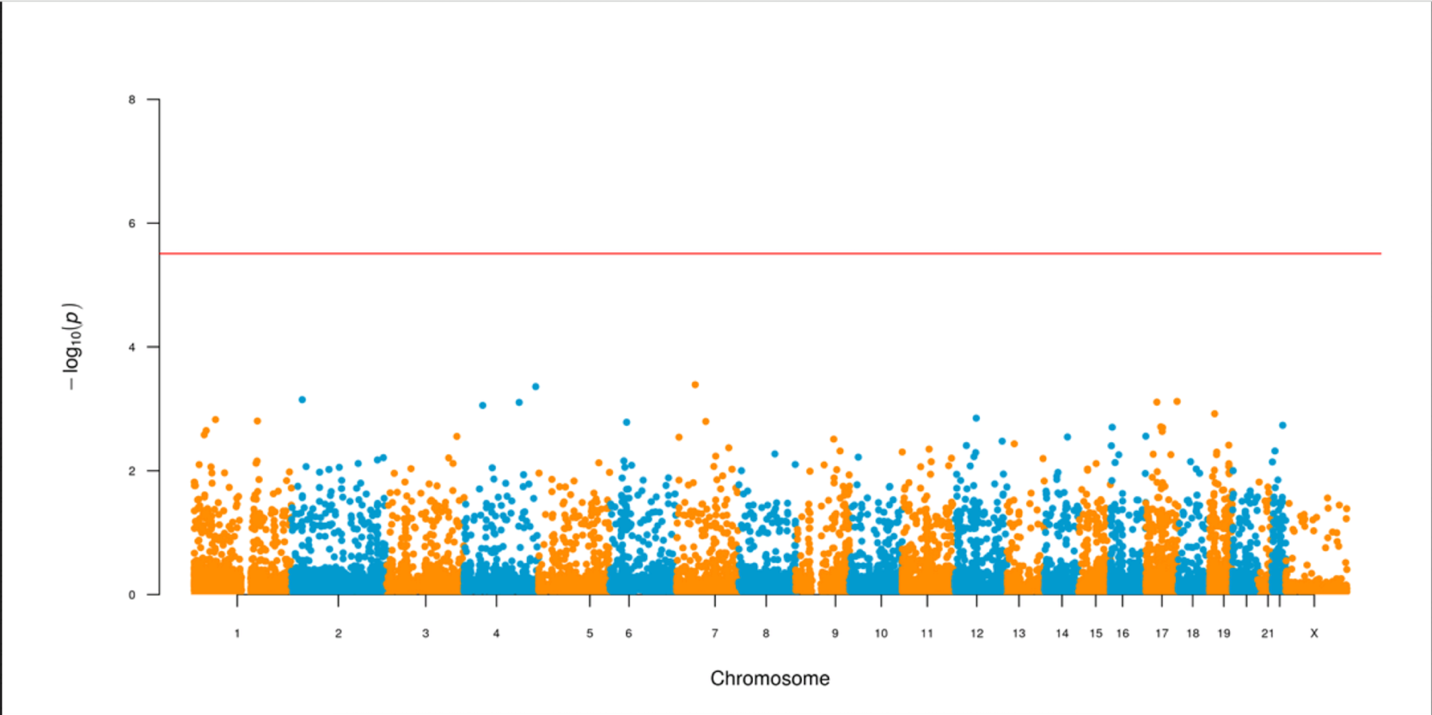

**Fig.S5** Manhattan style plot showing the results of the rare variant analysis, using gnomAD, for LoF variants with MAF < 0.001 (0.1%) in the HES-RR cohort, with the red line representing the -log_10_ of the P value (~*2.6 x 10^-6^*) that is the threshold for genome-wide significance. There is no enrichment of any LoF rare variant affecting the EEHTN phenotype in this cohort of 449 cases v 20852 controls.

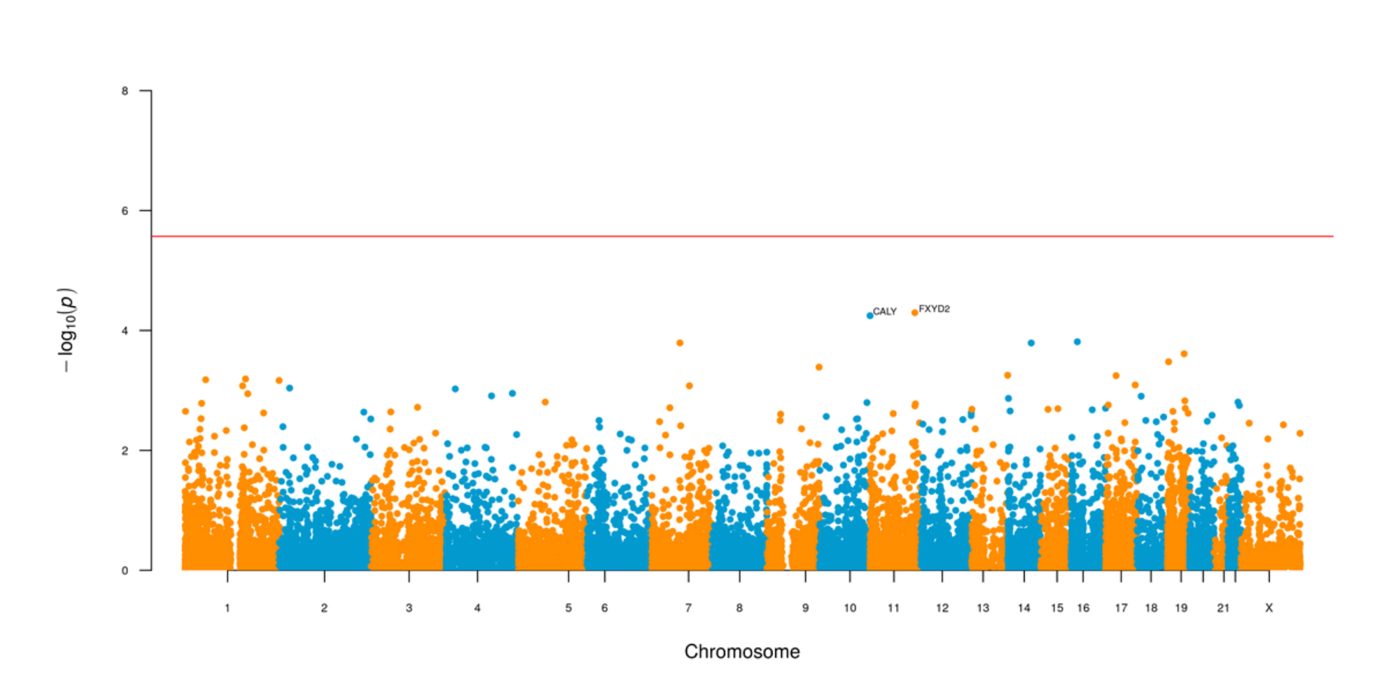

**Fig.S6** Manhattan style plot showing the results of the rare variant analysis, using gnomAD, for missense+ variants with MAF < 0.001 (0.1%) in the RR-EEHTN cohort, with the red line representing the -log_10_ of the P value (~*2.6 x 10^-6^*) that is the threshold for genome-wide significance. There is no enrichment of any missense+ rare variant affecting the EEHTN phenotype in this cohort of 449 cases v 20852 controls.

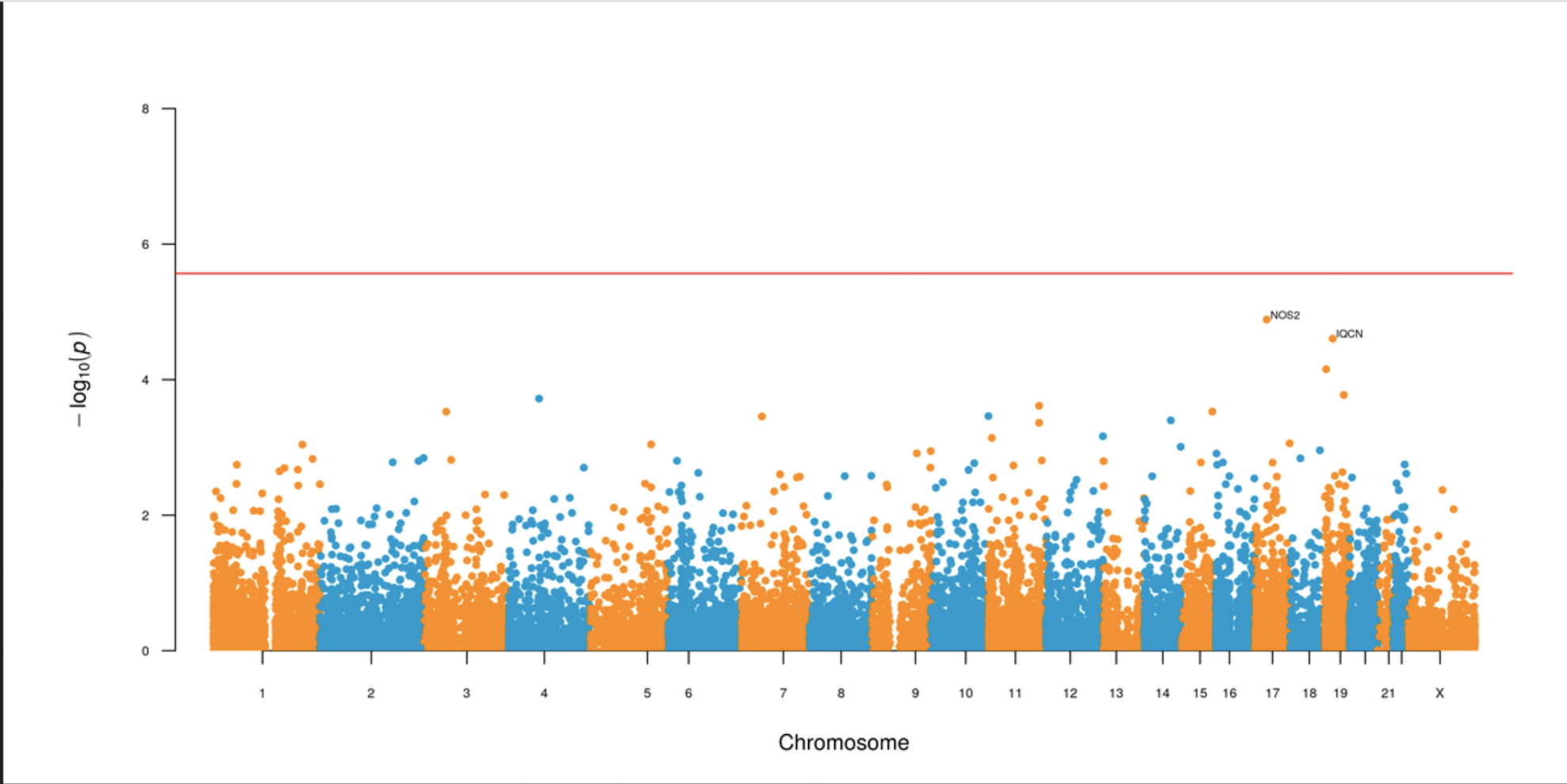

**Fig.S7** Manhattan style plot showing the results of the rare variant analysis, using gnomAD, for missense+ variants with MAF < 0.001 (0.1%) in the HES-EEHTN cohort, with the red line representing the -log_10_ of the P value (~*2.6 x 10^-6^*) that is the threshold for genome-wide significance. There is no enrichment of any missense+ rare variant affecting the EEHTN phenotype in this cohort of 901 cases v 20852 controls.

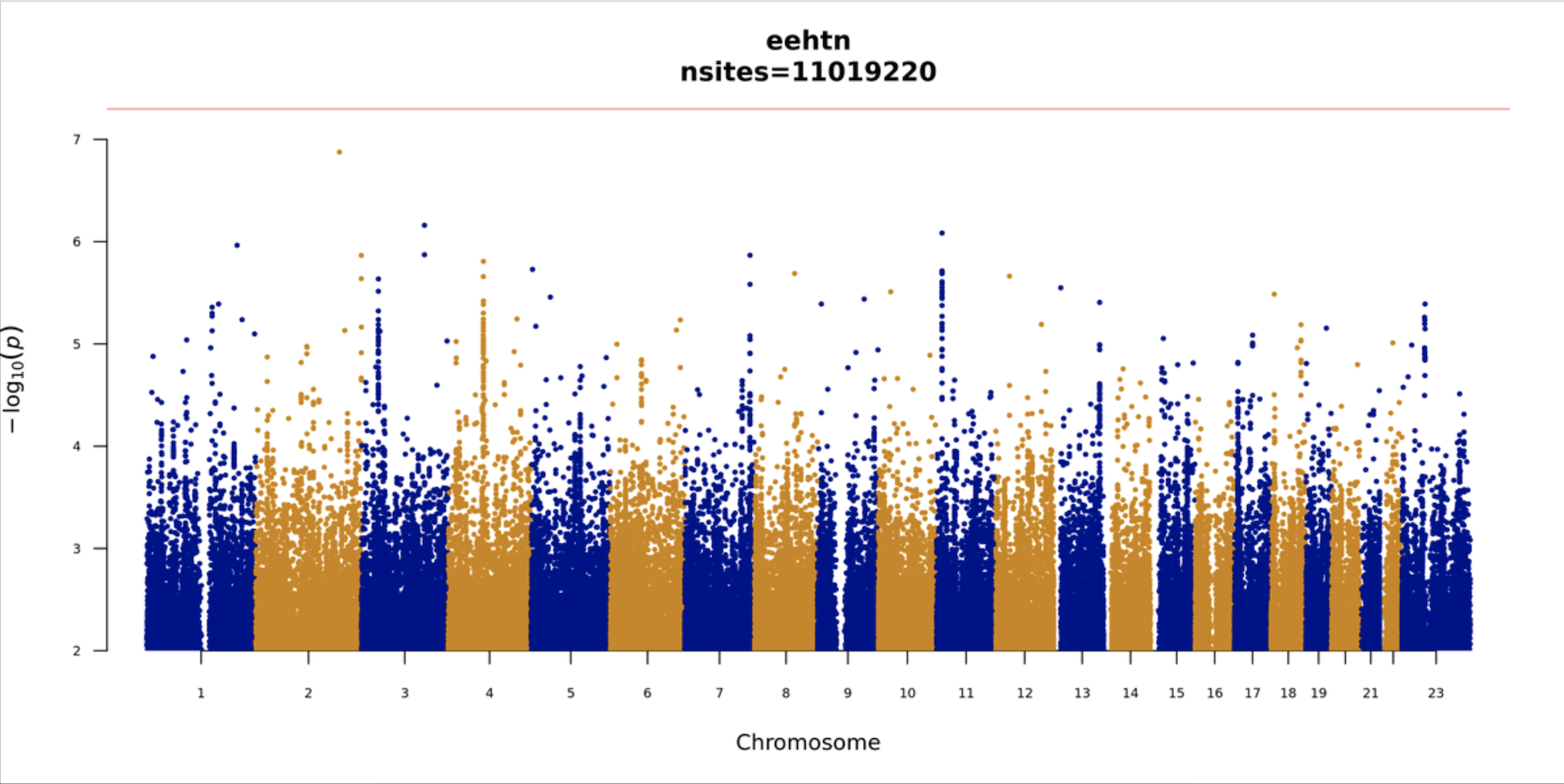

**Fig.S8** Manhattan plot showing the GWAS results of the initial EEHTN cohort, with the red line representing the -log_10_ of the P value that is the threshold for genome-wide significance. In this analysis 11019220 markers were tested, with MAF > 0.01 (1%). The statistical significance level for this analysis is approximately *4.54 x 10^-9^*, as represented by the red line. There is no enrichment of any SNV or indel affecting the EEHTN phenotype in this cohort of 179 cases v 20411 controls.

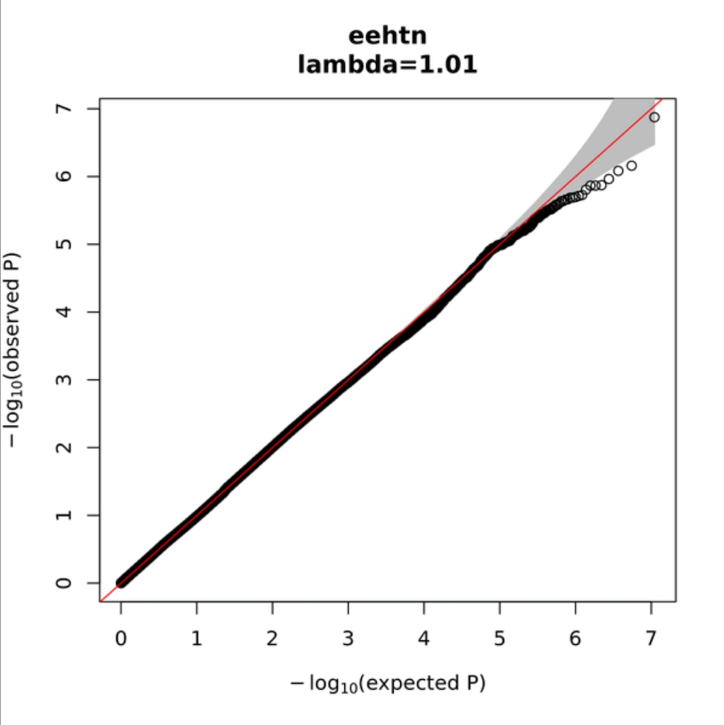

**Fig.S9** Quantile-Quantile plot (Q-Q plot) for the GWAS of the initial EEHTN cohort. The lambda of 1.01 suggests that population stratification has successfully been controlled for.

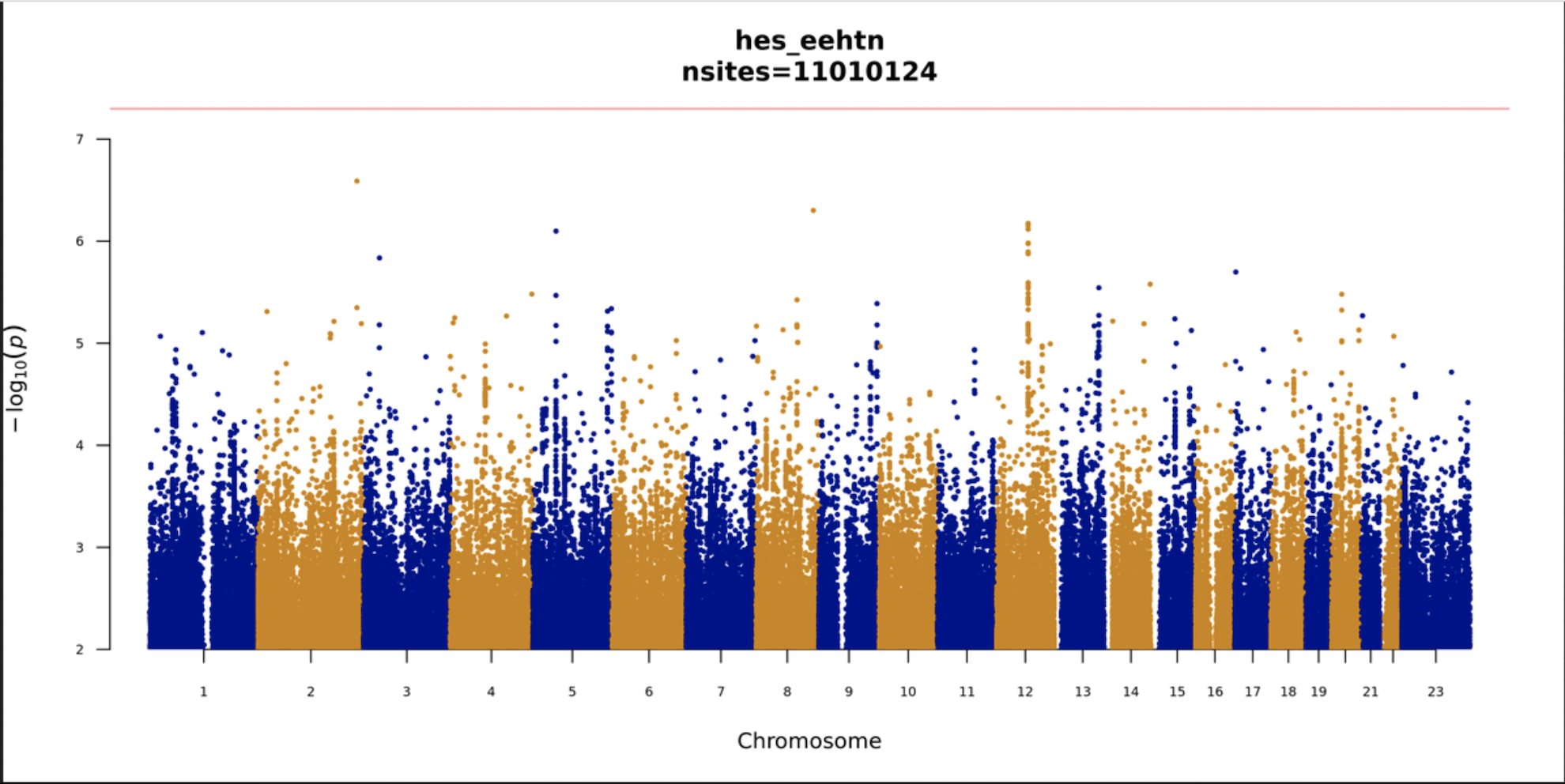

**Fig.S10** Manhattan plot showing the GWAS results of the HES-EEHTN cohort, with the red line representing the -log_10_ of the P value that is the threshold for genome-wide significance. In this analysis 11010124 markers were tested, with MAF > 0.01 (1%). The statistical significance level for this analysis is approximately *4.54 x 10^-9^*, as represented by the red line. There is no enrichment of any SNV or indel affecting the EEHTN phenotype in this cohort of 901 cases v 20852 controls, as none of the points lie above the red line which represents the genome-wide significance level.

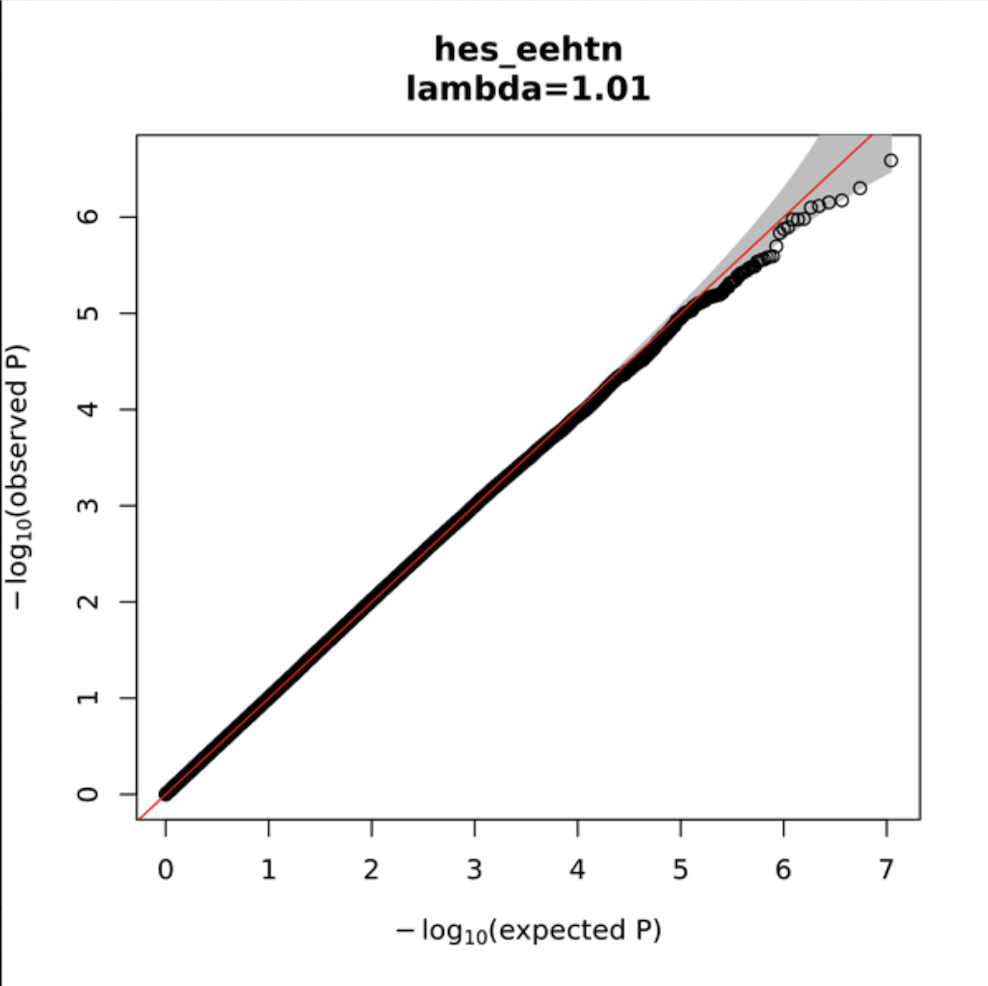

**Fig.S11** Quantile-Quantile plot (Q-Q plot) for the GWAS of the HES-EEHTN cohort. The lambda of 1.01 suggests that population stratification has successfully been controlled for.

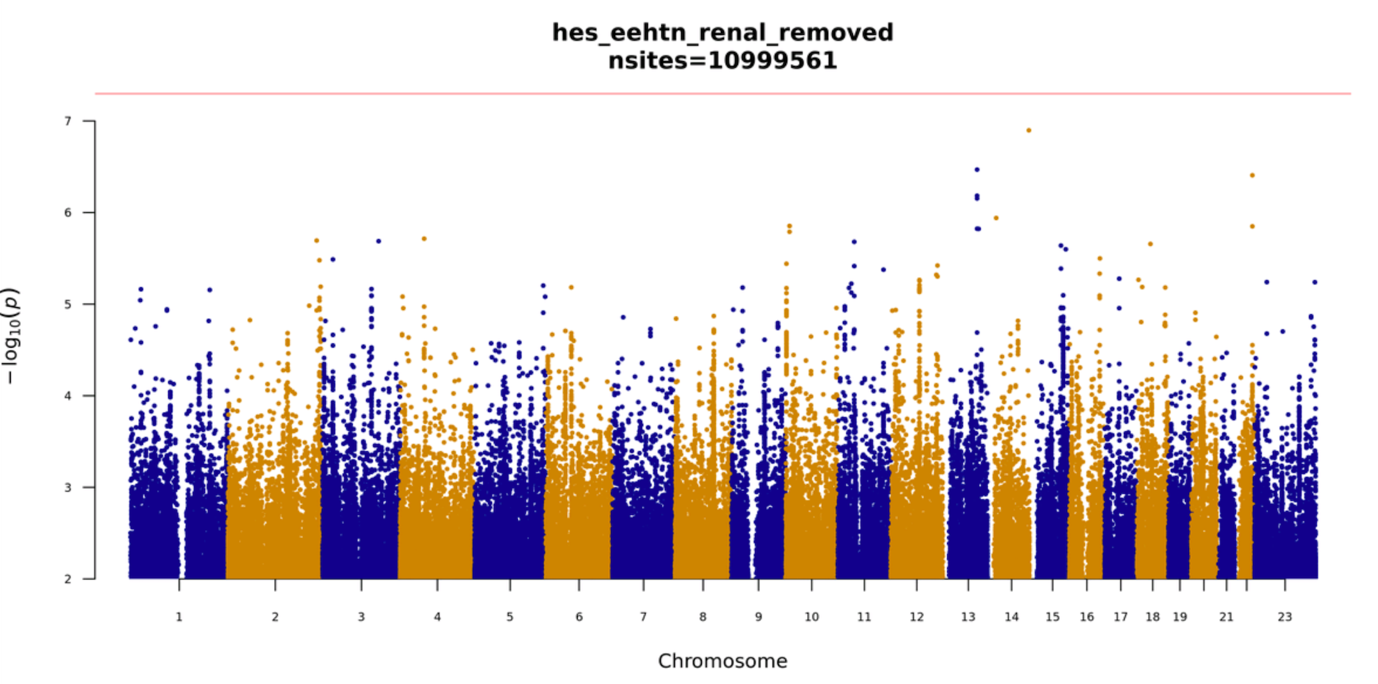

**Fig.S12** Manhattan plot showing the GWAS results of the RR-EEHTN cohort, with the red line representing the -log_10_ of the P value that is the threshold for genome-wide significance. In this analysis 10999561 markers were tested, with MAF > 0.01 (1%). The statistical significance level for this analysis is approximately *4.55 x 10^-9^*, as represented by the red line. There is no enrichment of any SNV or indel affecting the EEHTN phenotype in this cohort of 449 cases v 20852 controls, as none of the points lie above the red line which represents the genome-wide significance level.

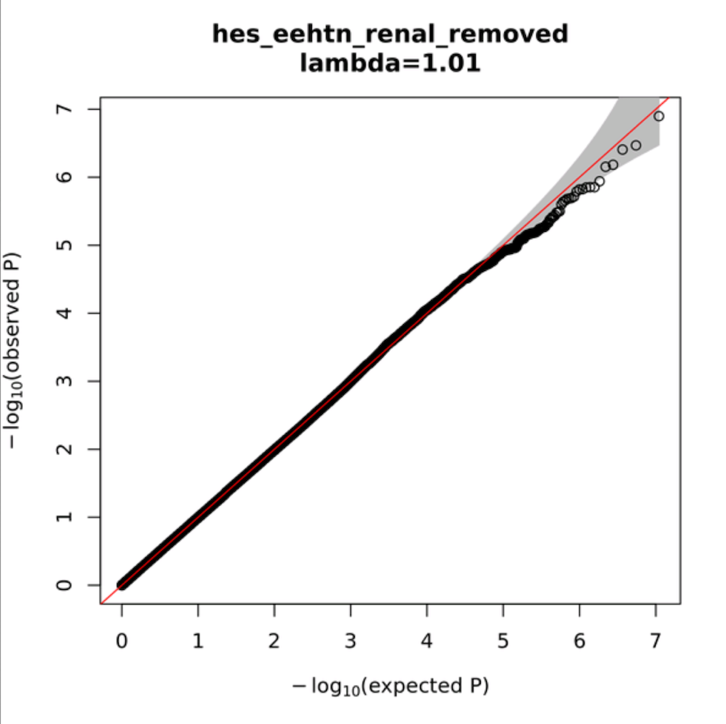

**Fig.S13** Quantile-Quantile plot (Q-Q plot) for the GWAS of the HES-RR cohort. The lambda of 1.01 suggests that population stratification has successfully been controlled for.

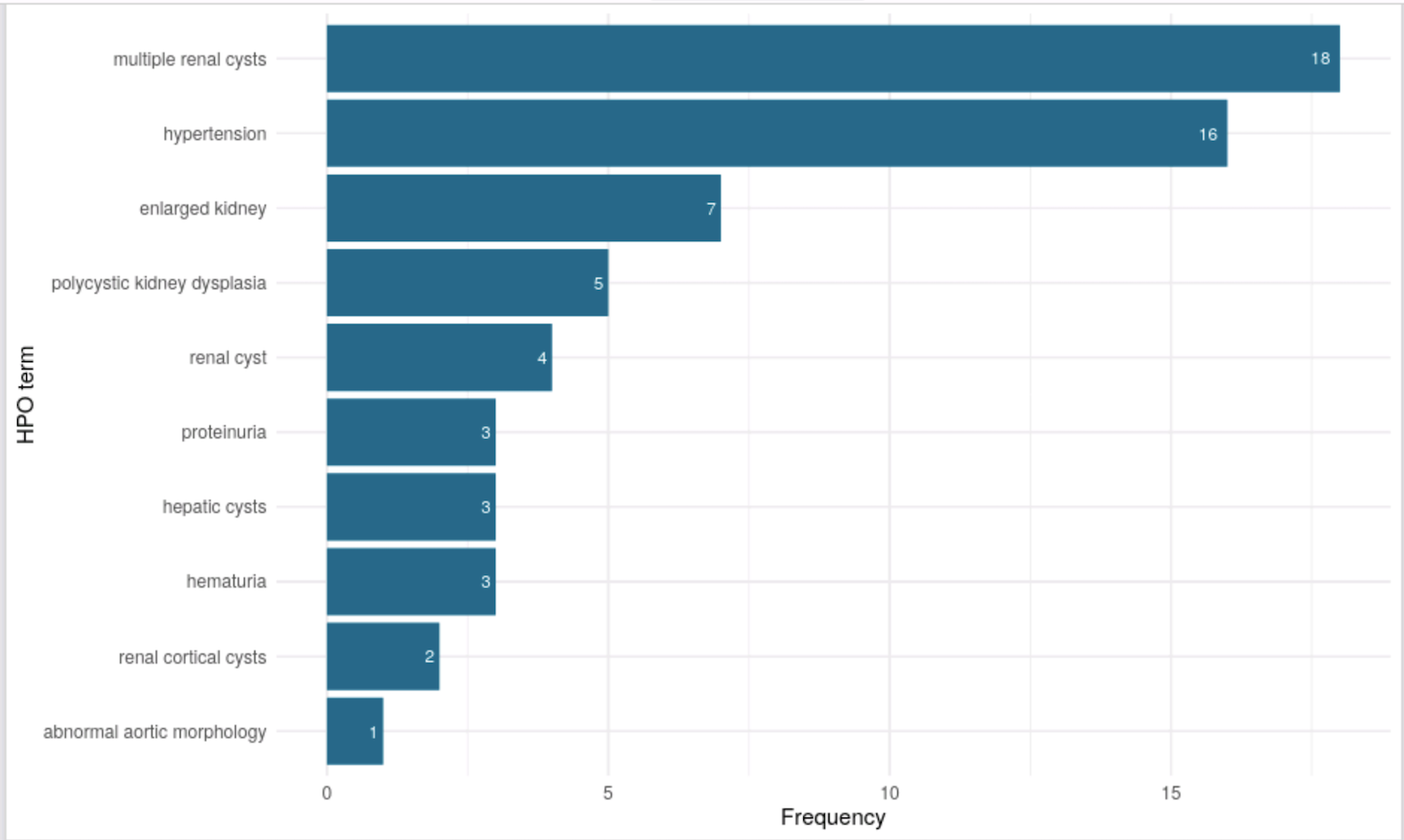

**Fig.S14** Graph showing the top 10 HPO codes in the cohort of patients with *PKD1* variants and the frequency of each one.
